# Anthropometrics: A forgotten gem in clinical assessment of obesity

**DOI:** 10.1101/2020.11.19.20234740

**Authors:** R Bhatti, U M. Warshow, M Joumaa, M ElSaban, F Nawaz, A H Khamis

## Abstract

**Background:** According to World Health Organization (WHO), United Arab Emirates (UAE) has one of the highest prevalence rates of obesity in the Middle East at 34%. There is a paralleled rise in the incidence of related metabolic conditions, particularly type 2 diabetes, metabolic syndrome and non-alcoholic fatty liver disease (NAFLD). Body mass index (BMI) alone is an insufficient marker of abdominal adiposity and addition of waist circumference (WC) can help to assess the cardiometabolic risk.

**Aim:** To study the prevalence of obesity related diseases in a multidisciplinary weight management program and determine the relationship to obesity anthropometric indices.

**Methods:** This is a cross-sectional study conducted at Mediclinic Parkview Hospital in Dubai, UAE. 308 patients have been evaluated from January 2019 until September 2019 as part of a multi-disciplinary weight management program. Key demographics, anthropometrics, and clinical data was analyzed using Statistical Package for Social Sciences software version 25 (SPSS Inc., Chicago, IL).

**Results:** Three hundred and eight patients taking part in the weight management program were studied. The population was constituted of 103 (33%) males and 205 (67%) females. The mean age was 41 years (±9.6) with a median BMI of 34.5 (±6.7) and 33.7 (±7.8) for males and females respectively. Mean waist circumference was 113.4 cm (±23.3) and 103.5 cm (±16.2), fat percent was 33.7% (±11.6) and 45 (±6.8), fat mass was 41 kg (±15.2) and 41.1 (±14.1), and visceral fat was 6.5 kg (±3.2) and 3.1 (±1.8), for males and females respectively. The population was heterogeneous with 38 nationalities. BMI strongly correlated with waist circumference (male; female, r=0.67; r=0.72) and visceral fat (male; female, r=0.89; r=0.78). Further, waist circumference was significantly associated with risk of diabetes, hypertension, and NAFLD.

**Conclusion:** The study has confirmed the high prevalence rates of obesity related diseases in a private hospital setting in a multinational cohort of obese patients. BMI and waist circumference are the most representative measures of obesity in our population and correlate with abdominal adiposity and obesity related diseases. Further studies will play a part in assessing the benefit of these measures during weight reduction interventions.

## Introduction

According to WHO prevalence of overweight is reported as 70.9% in females and 70.5% in males while the prevalence of obesity is reported as 41.2% in females and 31.6% in males in UAE.^1^The prevalence of overweight and obesity has increased worldwide as defined by BMI. The prevalence of overweight in Gulf Cooperation Council (GCC) (Bahrain, Kuwait, Qatar, Oman, Saudi Arabia and the UAE), in 2011, adults has been reported to be 48% amongst males and 35% amongst females, while the prevalence of obesity has been reported to be 24% amongst males and 40% amongst females.^2^ In UAE in particular, a study conducted at the national level between 1999 and 2000 reported prevalence rates of 40% and 30% for overweight and obesity, respectively, in Emirati and non-Emirati adults combined.^3^ In contrast, a more recent study reports a prevalence of 42% for overweight and 20% for obesity in 2012 among the same abovementioned population.^4^

There is a paralleled rise in the incidence of related metabolic conditions, particularly type 2 diabetes, metabolic syndrome and non-alcoholic fatty liver disease (NAFLD).^5^ However, BMI itself does not provide any information on body fat distribution, which may be more closely related to metabolic risk than BMI itself. Waist circumference is simple method in clinic practice to assess the abdominal adiposity. Waist circumference is strongly associated with cardiovascular mortality.^6^ Therefore; waist circumference should be used in conjunction with BMI to assess the metabolic risk according to World Health Organization report.^7^

The aim of our study was to look at anthropometric measures like WC, body fat percentage with BMI and their correlation with related metabolic conditions like diabetes, hypertension and NAFLD.

## Methods

### Study Design

This is a cross-sectional observational study conducted at Mediclinic Parkview Hospital, Dubai, and UAE between January-September 2019. 308 patients enrolled in the hospital’s multi-disciplinary weight management program were included in this study.

### Definitions

BMI was defined as weight divided by height squared (kg/m^2^).

Waist circumference (WC), was defined as measurement midway between the lowest rib and the iliac crest using a flexible tape.

Hip circumference (HC) was measured at the level of the greater trochanters to the nearest millimetre using a flexible tape.

Waist-to-hip ratios (WHR) were obtained by dividing waist circumference by hip circumference.

### Variables

According to World Health Organization (WHO) recommendations BMI is categorized as healthy weight (BMI 20–25), *overweight* (BMI 25–29.9) and *obese* (BMI≥30).^7^

Men with a waist circumference of <94, 94–101.9 and ≥ 102 cm were classified as normal weight, overweight and obese respectively, while women were classified in the same obesity categories on the basis of WC <80, 80–87.9 and ≥ 88 cm.

Men with WHR < 0.90, 0.90–0.99 and ≥ 1.0 were classified as normal weight, overweight or obese respectively, while women were classified in the same categories on the basis of WHR of < 0.80, 0.80 – 0.84 and ≥ 0.85.

Normal fat percentage for women is defined between 21-35%

Normal fat percentage man is defined between 8-24%

Normal visceral fat ratio for women is below 1.2

Normal visceral fat ratio for men is below 2.1

### Data collection

Data was collected from electronic medical records Bayanaty® (InterSystems IRIS, US) and Seca medical body composition analyzer 514(Seca®, Germany). Data collection was done in four categories: Demographic data, anthropometric measures, laboratory measurements, and clinical disease and risk factors status.

Demographic data included age, gender and nationality. Anthropometric measures included height, weight, BMI, fat mass, body fat percentage, and visceral fat mass, WC, HC and WHR. Laboratory measurements included glycated haemoglobin (HbA1c), renal function such as creatinine and estimated glomerular filtration rate (eGFR), liver function tests including AST and ALT, lipid profile including cholesterol, triglyceride, LDL, and HDL. Clinical variables included presence of diabetes, hypertension, polycystic ovarian syndrome, dyslipidemia and non-alcoholic fatty liver disease (NAFLD).

### The definition of metabolic risk factors

Four Metabolic syndrome components were included in the analysis: elevated BP (130 mmHg and/or diastolic blood pressure 85 mmHg or drug treatment for hypertension), HbA1c (6.5% or diabetes treatment), high TG concentration (150 mg/dl or 1.7 mmol/L or drug treatment for elevated triglycerides), and low HDL cholesterol.

(<40 mg/dl or 1.0 mmol/L in men and, 50 mg/dl or <1.3 mmol/L in women or drug treatment).

### Statistical methods

Data was entered in computer using IBM-SPSS for windows version 25.0 (SPSS Inc., Chicago, IL). Frequency tables and measure of percentage and measure of tendency and dispersion were performed as descriptive. Categorical variables were cross-tabulated to examine the independency between variables, for such variables the chi-square test or Fisher’s exact test as appropriate were used. Kolmogorov-Smirnov was used to test the normality of continuous variables. The Mann-Whitney test was used to compare the means between two groups if the normality was not confirmed while t-test was used for normal data per groups. A p-value of less than 0.05 was considered significant in all statistical analysis.

### Ethical Statement

Ethical approvals were taken from local Mediclinic Institutional Research Board; and Dubai Scientific Research Ethics Committee, Dubai Health Authority, Dubai, UAE.

## Results

A total of 308 patients were included in the study. 67% (n=205) were females. Mean age was 41.11 years. These patients represented 38 different nationalities. Basic demographics are shown in table 1.

**Table 1:** Basic demographics

|  |  |
| --- | --- |
| Total no of patients | 308 |
| Gender | No (%) |
| Male | 103 (33.4) |
| Female | 205 (66.6) |
| Age |  |
| Mean (SD) | 41.11 (9.6) |
| Nationality |  |
| Asia | 58 (19.1) |
| Arab | 94 (30.9) |
| Europe | 56 (18.4) |
| America | 18 (5.9) |
| Africa | 72 (23.7) |
| New Zealand and Australia | 6 (2) |

30% (n=91) were overweight and 70% (n=217) were obese according to WHO criteria using BMI and gender specific WC.

Table 2 represents gender specific general characteristics of patients. Males had higher weight, height, WC, BMI and visceral fat while females had higher % calculated fat which was statistically significant (p < 0.05)

**Table 2:** General characteristics of the sample

|  | Male (n=103) | Female (n=205) | p-value |
| --- | --- | --- | --- |
| Weight | 110.2 (23.8) | 88.9 (20.1) | < 0.001 |
| % fat calculated | 33.7 (11.6) | 41.8 (13.7) | < 0.001 |
| % fat | 75.4 (372.3) | 45.0 (6.8) | 0.43 |
| WC | 113.4 (23.3) | 103.5 (16.2) | < 0.01 |
| HC | 127.1 (12.2) | 124.2 (14.7) | 0.388 |
| W:H ratio | 0.93 (0.2) | 0.87 (0.1) | 0.078 |
| Height | 170.9 (35.5) | 155.4 (33.7) | < 0.001 |
| BMI | 34.5 (6.7) | 33.7 (7.8) | 0.006 |
| Fat mass | 41.0 (15.2) | 41.1 (14.1) | 0.86 |
| Visceral fat | 6.5 (3.2) | 3.1 (1.8) | <0.001 |

**Table 3:** Comparison of lab measurements between overweight cases and obese cases by gender

|  | Male |  |  | Female |  |  |
| --- | --- | --- | --- | --- | --- | --- |
|  | overweight | Obese | p-value | overweight | Obese | p-value |
| HbA1C | 5.62 (0.55) | 6.26 (1.62) | 0.326 | 5.75 (0.73) | 5.57 (0.95) | 0.15 |
| Creatinine | 94.82 (20.82) | 82.16<br>(20.33) | 0.231 | 62.35 (7.96) | 65.06 (7.98) | 0.203 |
| eGFR | 86.37 (19.68) | 93.68<br>(20.64) | 0.427 | 105.05<br>(11.7) | 98.83 (17.5) | 0.12 |
| AST | 35.75 (22.47) | 31.71<br>(22.58) | 0.65 | 21.86 (7.44) | 30.73<br>(29.21) | 0.74 |
| ALT | 31.25 (10.9) | 47.04<br>(38.69) | 0.451 | 21.94<br>(13.23) | 34.39<br>(29.78) | 0.049 |
| Cholesterol | 5.96 (0.29) | 9.46 (28.9) | 0.77 | 5.2 (1.8) | 10.4 (33.13) | 0.534 |
| TG | 1.24 (0.27) | 2.36 (1.33) | 0.01 | 1.14 (0.45) | 1.57 (0.79) | 0.067 |
| LDL | 3.44 (1.71) | 6.41 (20.67) | 0.73 | 3.59 (1.13) | 3.58 (0.86) | 0.978 |
| HDL | 2.35 (1.91) | 1.93 (6.18) | 0.735 | 1.48 (0.36) | 1.21 (0.3) | 0.004 |

Laboratory data was also compared between overweight and obese cases among different gender TG was found significantly higher among obese compared with overweight in males. ALT and HDL were found statistically significant between overweight and obese among females HDL was found less among obese while ALT was found higher among obese.

The proportion of diabetes was found significantly higher among obese male, while hypertension was found higher among obese female. Dyslipidaemia was higher among obese female. Levels of ALT as a surrogate for NAFLD were significantly higher in female obese patients (shown in Table 4)

**Table 4:**
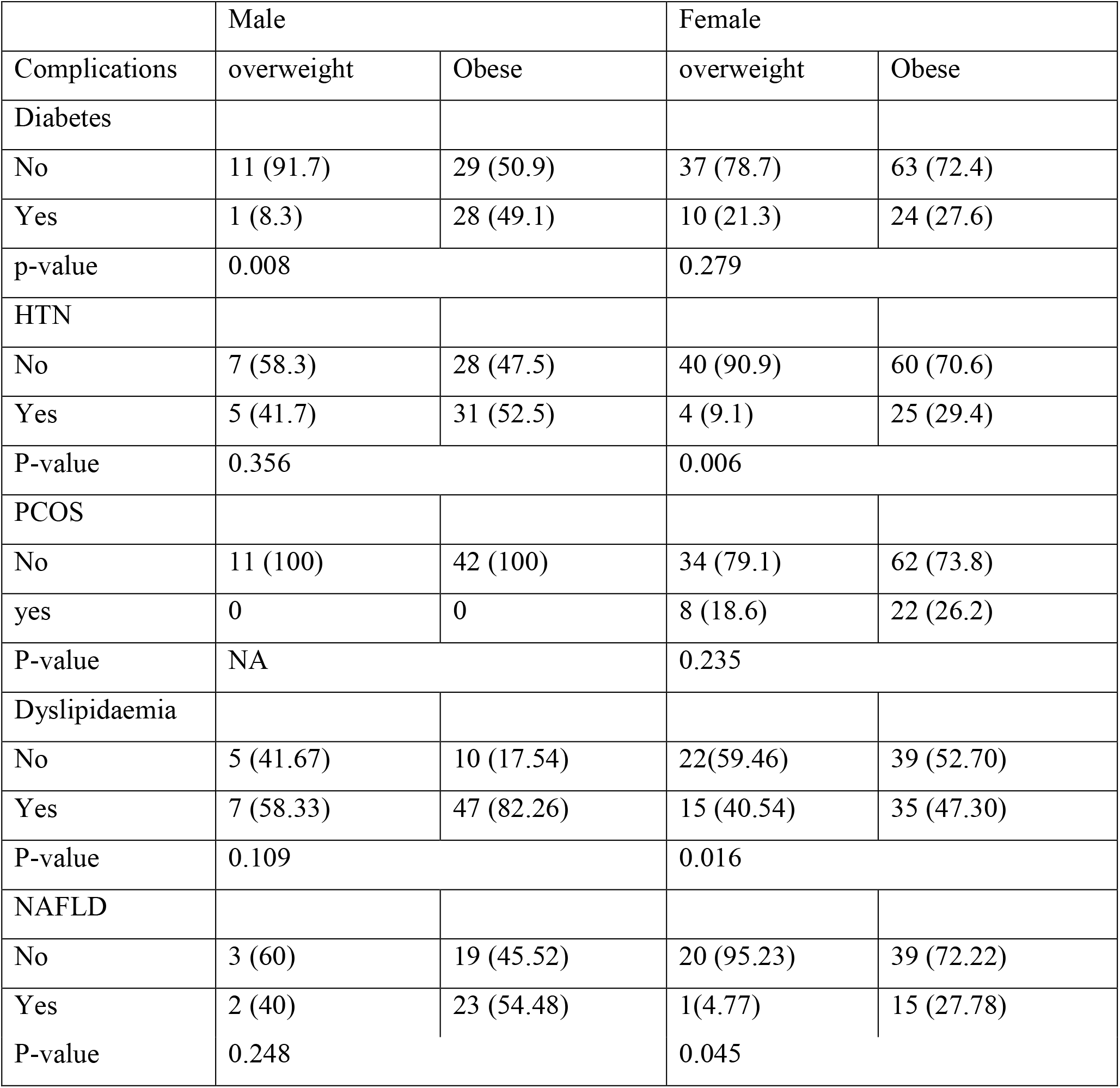
Comparison complications between overweight cases and obese cases by gender

|  | Male |  | Female |  |
| --- | --- | --- | --- | --- |
| Complications | overweight | Obese | overweight | Obese |
| Diabetes |  |  |  |  |
| No | 11 (91.7) | 29 (50.9) | 37 (78.7) | 63 (72.4) |
| Yes | 1 (8.3) | 28 (49.1) | 10 (21.3) | 24 (27.6) |
| p-value | 0.008 |  | 0.279 |  |
| HTN |  |  |  |  |
| No | 7 (58.3) | 28 (47.5) | 40 (90.9) | 60 (70.6) |
| Yes | 5 (41.7) | 31 (52.5) | 4 (9.1) | 25 (29.4) |
| P-value | 0.356 |  | 0.006 |  |
| PCOS |  |  |  |  |
| No | 11 (100) | 42 (100) | 34 (79.1) | 62 (73.8) |
| yes | 0 | 0 | 8 (18.6) | 22 (26.2) |
| P-value | NA |  | 0.235 |  |
| Dyslipidaemia |  |  |  |  |
| No | 5 (41.67) | 10 (17.54) | 22(59.46) | 39 (52.70) |
| Yes | 7 (58.33) | 47 (82.26) | 15 (40.54) | 35 (47.30) |
| P-value | 0.109 |  | 0.016 |  |
| NAFLD |  |  |  |  |
| No | 3 (60) | 19 (45.52) | 20 (95.23) | 39 (72.22) |
| Yes | 2 (40) | 23 (54.48) | 1(4.77) | 15 (27.78) |
| P-value | 0.248 |  | 0.045 |  |

Correlation between BMI, WC, HC, visceral fat, % of fat calculated and WHR was investigated using BMI and gender specific cutoffs for WC and other variables. The results are represented in figure 1.

**Figure 1:**
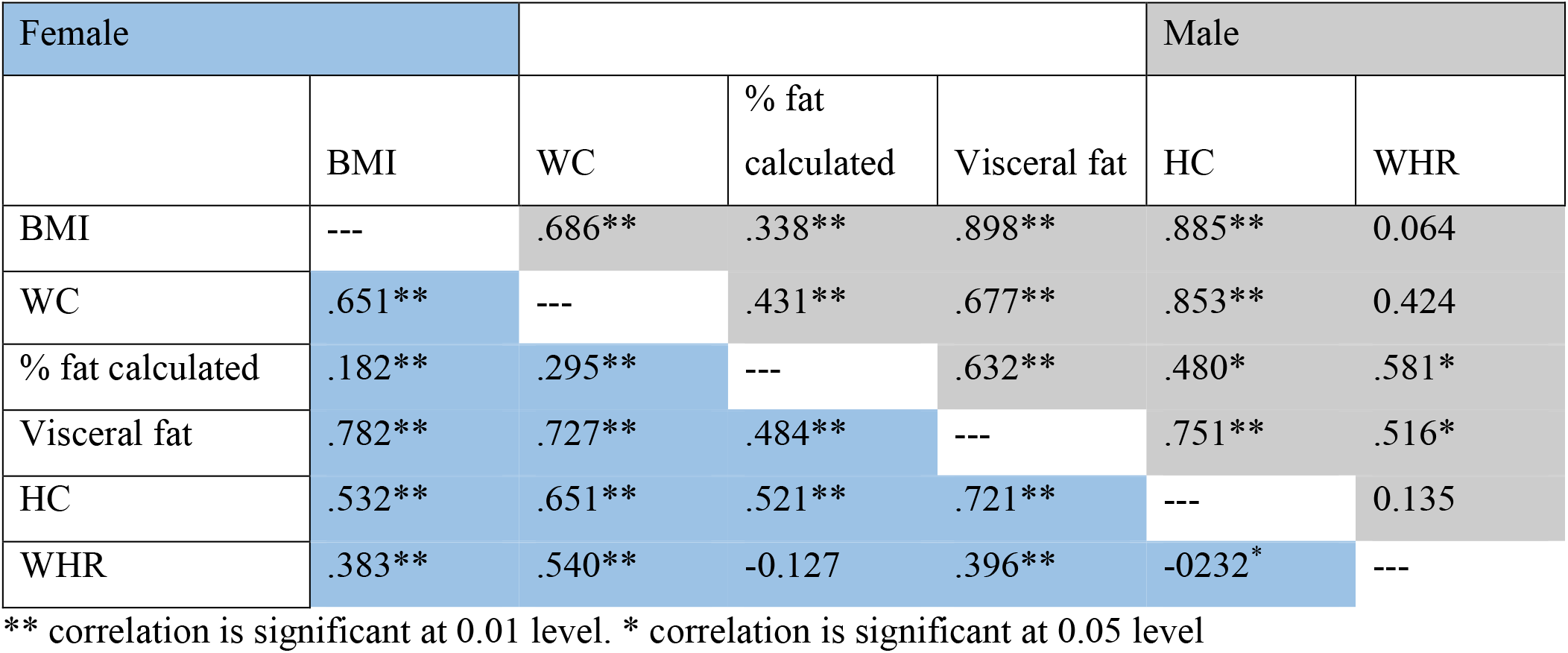
Matrix of correlation of the measurements of the indicators of anthropometrics of obesity.

BMI, WC and visceral fat showed strong significant correlation with each other while WHR showed weak correlation with other anthropometric measures.

## Discussion

In this study from a multidisciplinary weight management program in Dubai 30% were overweight and 70% were obese. According to WHO prevalence of obesity is reported as 34% in UAE in 2016 and our data is reflective of this high background prevalence.

Majority were female (67%) with participants from 38 different Nationalities as -Dubai is a multinational country with more than 200 nationalities.

BMI, WC and visceral fat show a strong significant correlation with each other. The correlation of WC to visceral fat establishes its benefit when combined with BMI and their correlation with each other suggests that measures of obesity based on these parameters will provide comparable information. Kamadieu et al demonstrated similar results in a Cameroon burden of diabetes baseline survey^8^. It is notable that increase in abdominal visceral adiposity is reflected by waist circumference and is related to increased cardio metabolic risk.^6^WHR has a weak correlation with other anthropometric measures in our cohort.

Our laboratory data showed that triglycerides were elevated in obese males. In obese women, ALT levels were significantly higher, as was prevalence of NAFLD. There was a statistically significant low HDL in obese females. Thus, our cohort reflected a gender difference in prevalence of obesity related conditions such hypertension, dyslipidemia and NAFLD.

The findings of our study has important implications on assessment of obesity in clinical practice as it reinforces the use of anthropometrics as indicators of obesity. The International atherosclerosis society (IAS) and international chair on cardiometabolic risk (ICCR) working group also published a consensus statement on visceral obesity in March 2020. It also recommended use of WC as a critically important target for reducing adverse health risks for both men and women.^9^ Recent Canadian guidelines for obesity in adults also recommend measurement of waist circumference in addition to BMI to identify individuals with increased visceral adiposity and adiposity related health risks.^10^

The limitations of our study include small sample size. More studies are needed to see the implications of anthropometrics on clinical outcomes of different weight management interventions.

## Conclusion

According to findings of our study, there is strong correlation between BMI, visceral fat and waist circumference. It highlights the importance of using anthropometrics such as waist circumference as measure of obesity in addition to BMI and it is easy and inexpensive clinical tool.

## Data Availability

All data is available on request

## Abbreviations

(BMI): Body mass index
(WC): Waist circumference
(HC): Hip circumference
(WHR): Waist hip ratio
(NAFLD): Non-alcoholic fatty liver disease

